# Accurate Detection of Lead Malfunction From ECG-derived Bipolar Pacing Stimulus Amplitude

**DOI:** 10.1101/2024.01.12.24301251

**Authors:** Mary Pelling, Michael S. Lloyd, Rand Ibrahim, Mikhael F El-Chami, Shahriar Iravanian

**Author notes:** **Corresponding author:** Michael S. Lloyd FHRS, 1364 Clifton Rd NE Suite F424, Atlanta, GA 30322. Mary Pelling and Michael Lloyd are joint first authors. **Authorship contribution:** Pelling, writing; Lloyd, concept and writing; Ibrahim, Figures and data analysis; El-Chami, data acquisition; Iravanian, writing.

## Abstract

**Background:** One of the most common modes of lead failure is outer insulation breach which may result in myopotential noise and device malfunction. “Pseudo-unipolarization” of bipolar pacing stimuli, as observed from a routine 12-lead ECG has been observed with insulation breaches. We sought to characterize this ECG finding to detect lead this type of lead malfunction.

**Methods:** 138 transvenous leads were analyzed (88 with known malfunction and 50 normal leads). The highest amplitude (any of 12-leads on standard ECG, 10mm/mV, GE Marquette) of a bipolar pacing stimulus on ECG was recorded and compared to a control dataset of newly implanted leads. An ROC curve for maximum ECG bipolar pacing stimulus amplitude was generated for prediction of lead functional status (normal vs malfunction).

**Results:** The cohort (49% females, 34% non-white) had an average age of 67 ± 16 years at implant. The malfunction group consisted of 61% RA and 39% RV leads with mean pacing output 2.74V at 0.5ms. There was a significant difference in ECG bipolar stimulus amplitudes at time of identification of failure (15.06 ± 13.533mm or 7.89 ± 7.56mm per V, p<0.001) compared to those of normal leads (2.54 ± 1.265mm or 0.86 ± 0.41mm per V). An EKG stimulus amplitude cut-off at 3.5mm for the prediction of this type of lead malfunction demonstrated a sensitivity of 86.4% and a specificity of 76%. When normalized for programmed stimulus output, a cutoff of 5mm/V demonstrated a sensitivity of 91% and a specificity of 92% (AUC 0.967 95% CI 0.938-0.996).

**Conclusion:** For a given output, the maximum amplitude of a bipolar pacing stimulus on ECG is significantly lower in normal functioning leads compared to those with known malfunction due to insulation breach. This simply-derived variable demonstrated good accuracy at identifying this lead failure due to insulation breach and exposed electrodes.

## Introduction

Transvenous lead failure can occur due to the breakdown of any of the lead components (insulation, conductors, connectors, terminal pins, electrodes, and coils) [1]. Over 10 years, lead failure rates have been reported as high as 20% in adults and 60% in children - with the most common mode of failure being an insulation breach [2]. Insulation breaches most commonly occur on the outside of the lead in the pocket or near the clavicle risking conductor exposure to tissue. This can have serious clinical consequences including inappropriate inhibition of pacing, short-circuit diversion of defibrillation, and oversensing of noise causing inappropriate defibrillation. Our group and others have reported an increased incidence of insulation breaches manifesting as low impedance and high frequency, low amplitude noise presumed from myopotentials - particularly in one lead family [3–5]. Changes in pacing impedance have historically been used to indicate lead insulation breakdown. Impedance values have not been reliable because, while most leads with insulation breaches may have impedance changes, the absolute value of the impedance remains in the normal range. This is because, even in the event of an alternate pathway from exposed conductor, the main determinant of total impedance is at the lead tip-myocardial interface [6].

A current generation surface 12-lead electrocardiogram (ECG) can reliably detect pacing stimuli. The most commonly used model in the US (10mm/mV, GE Marquette) has a digital sampling rate of 4000hz, a frequency response of [−3] dB @ 0.01 to 150 Hz and displays amplitudes with an accuracy of +/−5% and 200dpi resolution at standard speeds. Pacing stimuli with standard outputs, e.g., 0.4-10 volts and.2 - 1.5ms have frequency content that are reliably detected by these amplifiers, irrespective of automated pacing detection algorithms, used to artificially “tag” pacing stimuli.

Pacing stimuli can either be unipolar or bipolar. In unipolar pacing configuration the current path consists of the tip electrode to the impulse generator and results in a much higher-amplitude deflection on current-generation ECG machines from the larger current path in the body. In contrast, a bipolar pacing configuration current path travels between ring and tip electrodes resulting in much smaller amplitude deflections. (Figure 1)

**Figure 1.**
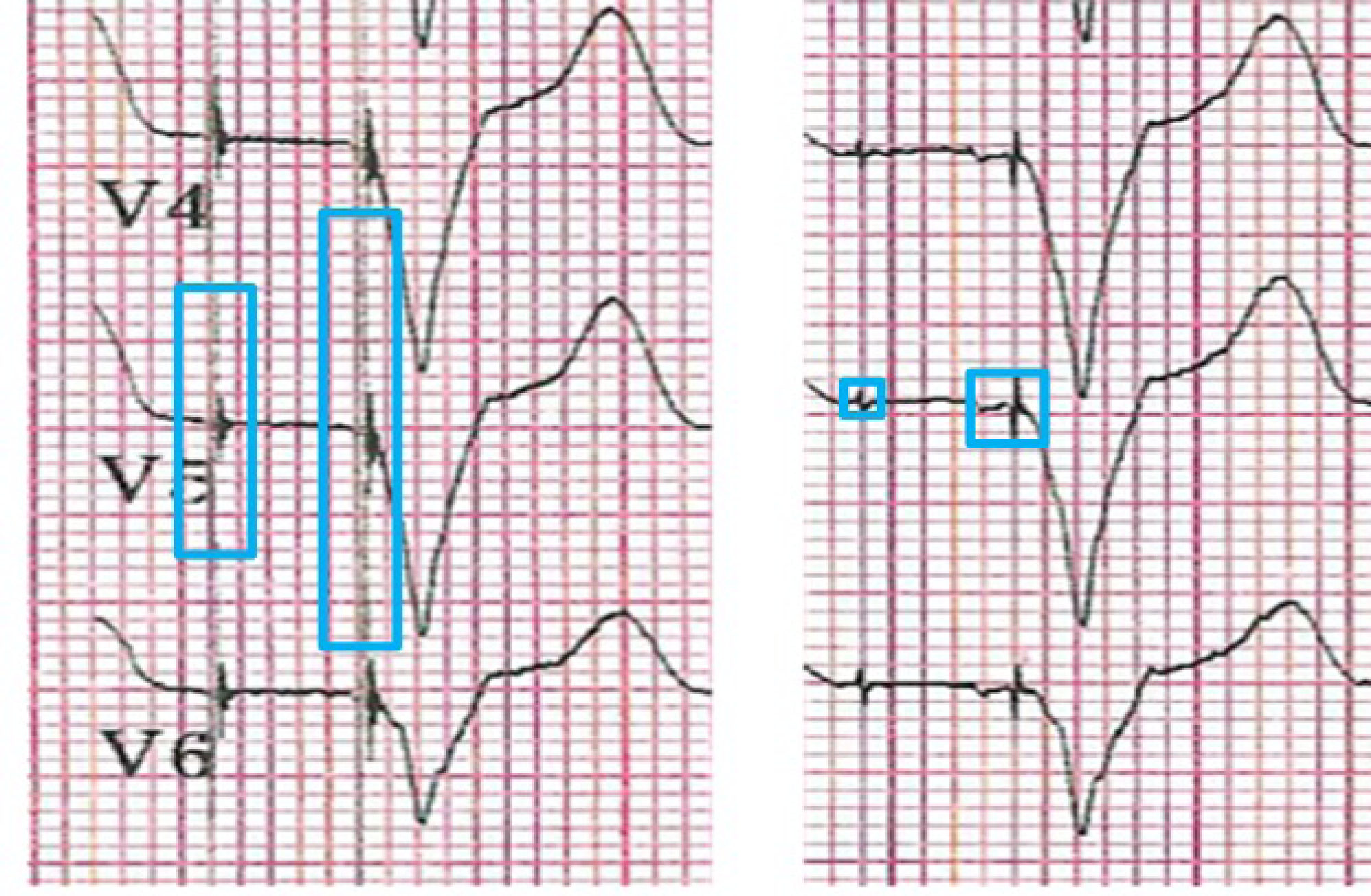
ECG amplitudes of unipolar pacing (left) and bipolar pacing (right) of same patient at identical output (3 V @ 0.5 ms).

It has been observed that surface ECG bipolar pacing amplitudes are larger in leads with insulation breach. The parallel current path of an exposed electrode at the site of breach and pacing tip has larger inter-electrode distance and results in a form of “pseudo-unipolarization.” The amplitude of this pacing stimulus may correlate more specifically with defects in lead insulation integrity. We hypothesized that a 12-lead ECG bipolar pacing stimulus amplitude would serve as a means of detection for this common type of lead malfunction.

## Methods

Institutional board review approval was received prior to the study and informed consent was waived by the board due to study design. Patients with CIED and transvenous lead implantation with pacing captured on ECG within the Emory Healthcare system were analyzed. Patients were included if they underwent lead revision for known lead malfunction. They must have had current evidence of insulation breakdown, including electrogram lead noise noted on device interrogation and/or a significant drop in lead impedance requiring device reprogramming or lead revision. The control cohort consisted of patients with newly implanted leads having had ECGs with pacing stimuli on the day of implant. While CRT devices were included in the analysis, only bipolar (ring to tip) stimuli from RA and RV pacing stimuli were used.

The highest amplitude (any of 12-leads on standard ECG, 10mm/mV, GE Marquette) of the bipolar pacing stimulus on ECG was recorded. The absolute height (below and above ECG isoelectric baseline) was used for this value. ECG bipolar stimulus amplitude was compared among the two groups. Additionally, a subset of the malfunction group whose pacing outputs were known at the time of ECG were normalized according to programmed pacing output to derive mm/V values and compared to normalized values in the normal group. Chi-squared analysis was used to compare categorical variables and independent t-test was applied for continuous values. The data were used to construct ROC curves for maximum ECG bipolar pacing stimulus amplitude according to lead functional status (normal versus malfunction). Routine demographic data including gender, age, race, and body mass index was evaluated. Device and lead information were also noted, including device type, manufacturer, model, lead chamber, time to lead breakdown in malfunction group, percentage pacing, lead impedance, and lead output.

## Results

### Patient Characteristics

A total of 138 transvenous leads were included in the study. 50 leads in 38 patients with pacing noted at time of device implant served as the normal control group. 85 patients with lead insulation breakdown defined as above served as the malfunction group with 3 patients exhibiting malfunction on both atrial and ventricular leads for a total of 88 leads for analysis.

Table 1 includes patient demographics of those included in the study as well as a comparison between malfunction and control groups. The average body mass index (BMI), gender, and race between the control and malfunction patients were similar, however, patients in the control group were significantly older (74 ± 13 years vs. 64 ± 16 years, p=<0.001).

**Table 1.** Patient characteristics.

|  | **Overall**<br>*n* = 138 | **Control**<br>*n* = 50 | **Malfunction**<br>*n* = 88 | **P value** |
| --- | --- | --- | --- | --- |
| **Age** | 67 ± 16 | 74 ± 13 | 64 ± 16 | **< 0.001** |
| **BMI** | 27.8 ± 6.5 | 27.0 ± 4.5 | 28.1 ± 7.3 | 0.322 |
| **Gender (Male)** | 61 (50%) | 17 (45%) | 44 (52%) | 0.471 |
| **Race** |  |  |  | 0.293 |
| Caucasian/ White | 78 (63%) | 22 (58%) | 56 (66%) |  |
| African American | 38 (31%) | 12 (32%) | 26 (31%) |  |
| Asian | 1 (1%) | 1 (2%) | 0 (0%) |  |
| Unknown | 5 (4%) | 3 (6%) | 2 (2%) |  |
| American Indian or Alaskan | 1 (1%) | 0 (0%) | 1 (1%) |  |

### Device and Lead Characteristics

The study populations significantly varied in their composition of device type and manufacturer as displayed in Table 2. In the control group, the majority of devices were dual chamber pacemakers (70%) and CIEDs made by Medtronic (MDT) (68%). The control group included 3 CIEDs made by Abbott (ABT, formerly St. Jude Medical). In comparison, in the malfunction group 47% of the devices were dual chamber pacemakers and 30% CRT-Ds (p=0.008, control vs. malfunction). The malfunction group had significantly more CIEDs made by ABT (45%) followed by MDT (30%) and 20% Boston Scientific (BS) (p=<0.001, control vs. malfunction).

**Table 2.**
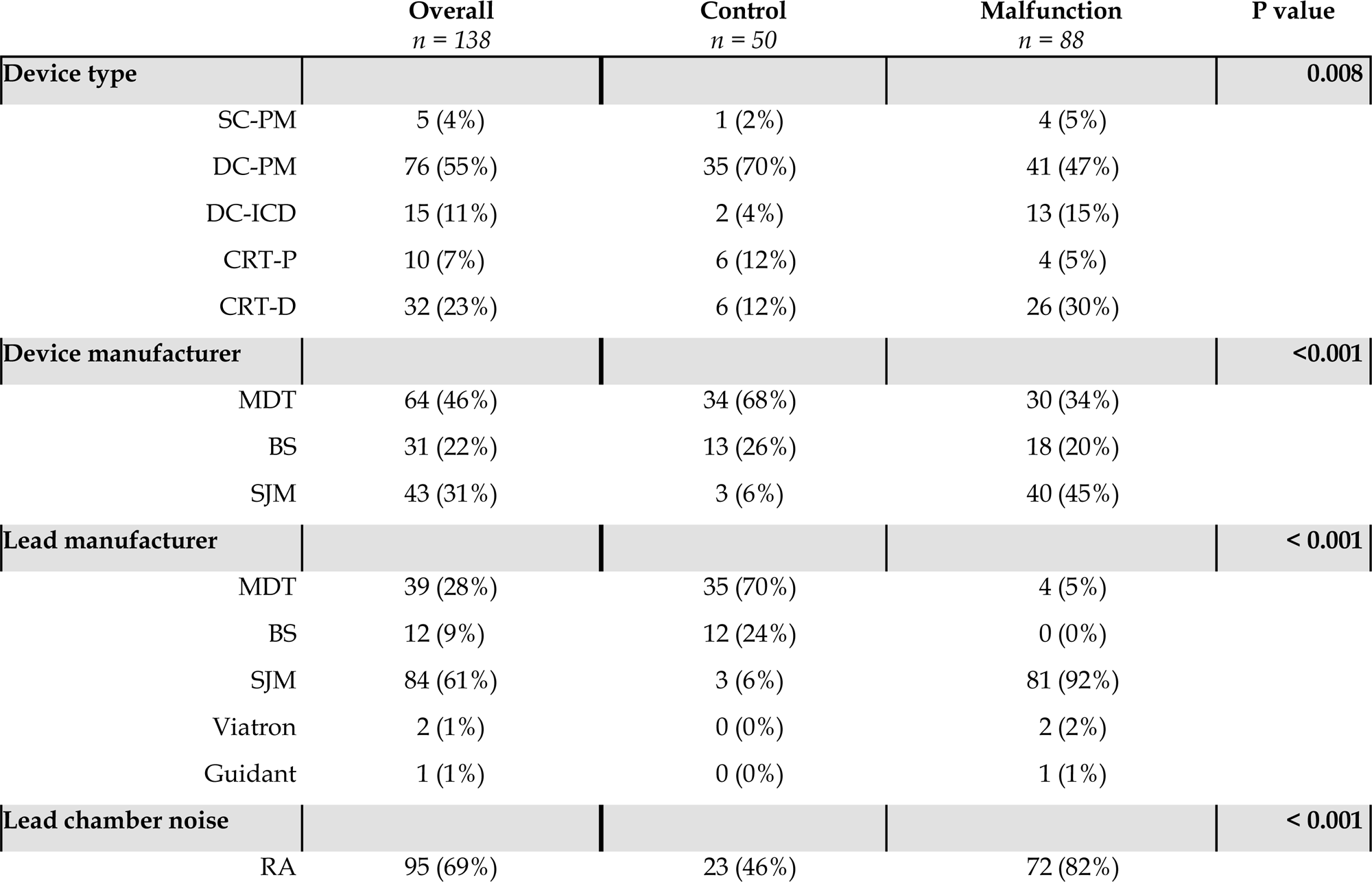
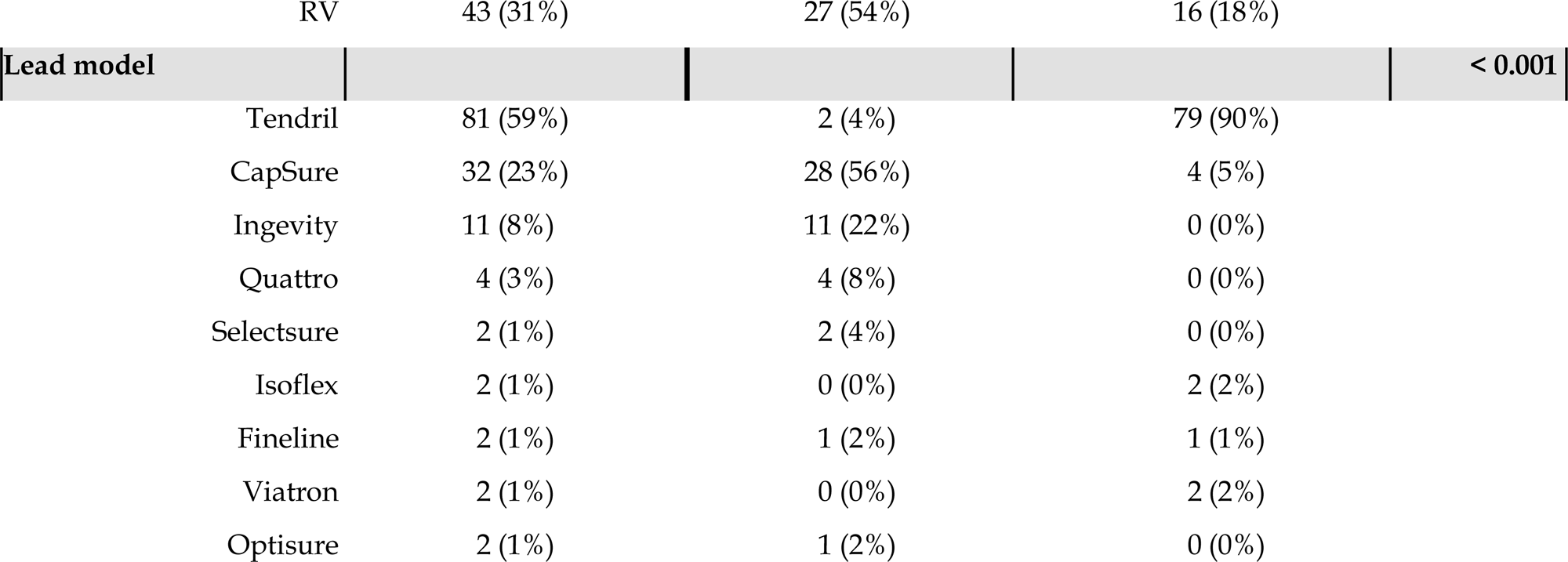
Device and lead characteristics.

The location and type of leads analyzed among each study population are shown in Table 2. In the control group leads were somewhat evenly distributed with 46% of the leads located in the right atrium (RA) and 54% in the right ventricle (RV). Whereas in the malfunction group, 82% of the leads were in the RA and only 18% in the RV (p=<0.001, control vs. malfunction). Most control group leads were made by MDT (70%) while 92% of the malfunction leads were made by ABT (p=<0.001) with 90% coming from the Tendril family of leads.

Lead diagnostics were compared among the two study groups, shown in Table 3. Atrial lead impedance was modestly, but significantly lower in the malfunctioned leads compared to the control group (409.1 ± 127.2 Ohms vs. 516.1 ± 119.5 Ohms, p=0.001) but still remained in normal range. No statistical difference was found among RV lead impedances between the proven malfunctioning leads and the control (510.3 ± 334 Ohms vs 550.6 ± 103.3 Ohm, p = 0.64). Mean programmed device outputs were significantly higher in the control group (RA: 2.18 ± 0.85 Volts [V] vs 2.93 ± 0.81 V, p = 0.001; RV: 1.98 ± 0.81 V vs 3.12 ± 0.85 V, p = 0.001), which is to be expected as thresholds are purposefully programmed high during the peri-implant period and limited sample size data was available for this analysis.

**Table 3.** ECG stimulus amplitude measurements.

|  | Overall<br>*n* = 138 | Control<br>*n* = 50 | Malfunction<br>*n* = 88 | P Value |
| --- | --- | --- | --- | --- |
| ECG Amplitude (Overall) | 10.5 ± 12.4 | 2.54 ± 1.26 | 15.06 ± 13.53 | <0.001 |
| ECG Amplitude (A) | 12.0 ± 12.7 | 2.5 ± 1.2 | 15.1 ± 13.2 | <0.001 |
| ECG Amplitude (V) | 7.1 ± 11.0 | 2.6 ± 1.4 | 14.8 ± 15.4 | 0.006 |

### ECG Amplitude Analysis

There was a significant difference in ECG bipolar stimulus amplitudes between study populations, detailed in Table 4. At time of failure, average bipolar stimuli amplitude of 15.1 ± 13.5 mm was significantly higher than normal leads with amplitude of 2.5 ± 1.3mm (p<0.001). Similarly, comparing atrial malfunctioned leads with controls showed significantly higher EKG stimulus (15.1 ± 13.2 mm vs. 2.5 ± 1.2 mm, p=0.001) as well as 5 ventricular malfunctioning leads (14.8 ± 15.4 mm vs. 2.6 ± 1.4 mm, p=0.006, Figure 2). This was despite a lower overall stimulus output in the experimental group compared to control.

**Figure 2.**
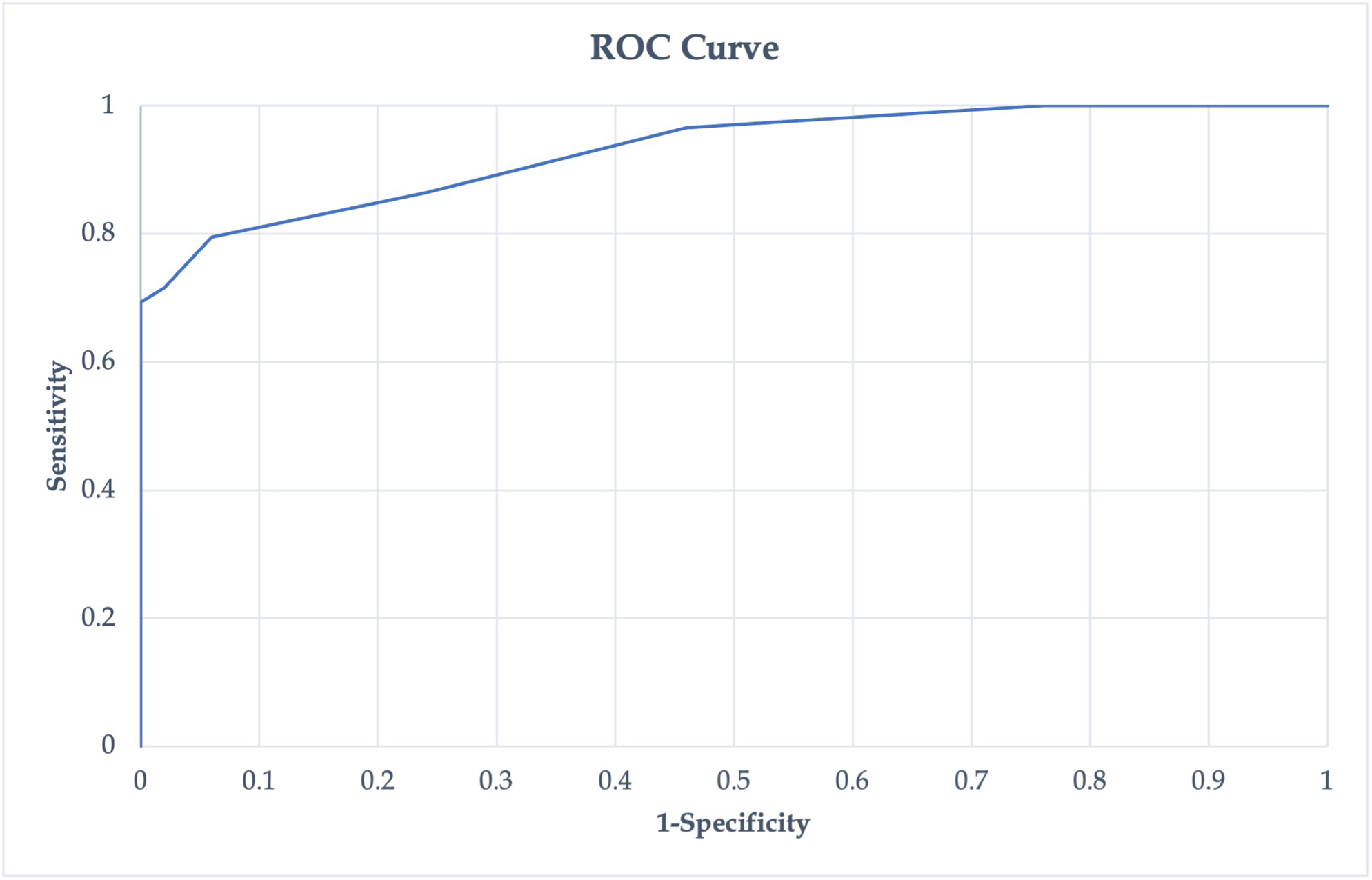
ECG stimulus amplitude analysis comparing malfunctional leads with functional (control) leads.

**Table 4.**
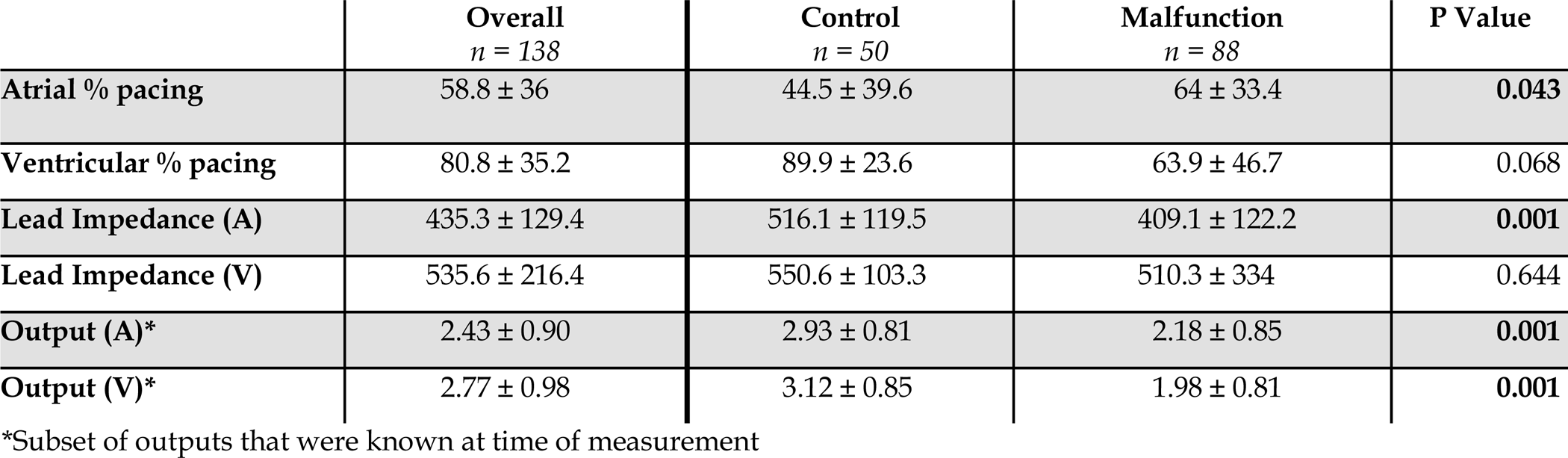
Lead function measurements.

|  | Overall<br>*n* = 138 | Control<br>*n* = 50 | Malfunction<br>*n* = 88 | P Value |
| --- | --- | --- | --- | --- |
| Atrial % pacing | 58.8 ± 36 | 44.5 ± 39.6 | 64 ± 33.4 | 0.043 |
| Ventricular % pacing | 80.8 ± 35.2 | 89.9 ± 23.6 | 63.9 ± 46.7 | 0.068 |
| Lead Impedance (A) | 435.3 ± 129.4 | 516.1 ± 119.5 | 409.1 ± 122.2 | 0.001 |
| Lead Impedance (V) | 535.6 ± 216.4 | 550.6 ± 103.3 | 510.3 ± 334 | 0.644 |
| Output (A)* | 2.43 ± 0.90 | 2.93 ± 0.81 | 2.18 ± 0.85 | 0.001 |
| Output (V)* | 2.77 ± 0.98 | 3.12 ± 0.85 | 1.98 ± 0.81 | 0.001 |

\*Subset of outputs that were known at time of measurement

A subset of patients in the malfunction group (58 leads) whose pacing outputs were certain at time of EKG were taken to compare normalized stimulus amplitudes. At time of failure, average bipolar stimuli amplitude of 7.89 ± 7.56 millimeters (mm) per V was significantly higher than normal leads with amplitude of 0.86 ± 0.41 mm per V (p<0.001). Atrial malfunctioned leads with controls showed significantly higher ECG stimulus (7.46 ± 6.88 mm per V vs 0.88 ± 0.44 mm per V, p < 0.001) as well as ventricular malfunctioning leads (7.94 ± 7.91 mm per V vs 0.85 ± 0.41 mm per V, p = 0.006) seen in Figure 2.

Receiver operating characteristic (ROC) curves for absolute and normalized maximum ECG bipolar pacing stimulus amplitudes were generated for the prediction of normal versus abnormal lead functional status shown in Figure 3. ROC amplitude displayed area under curve of 0.93 and 95% confidence interval 0.891-0.969. Selecting an EKG stimulus amplitude cutoff at 3.5 mm for the prediction of lead malfunction demonstrated a sensitivity of 86.4% and a specificity of 76%. For normalized data, ROC amplitude displayed area under curve of 0.967 and 95% confidence interval 0.938-0.996. Selecting an ECG stimulus amplitude cutoff at 5 mm per V for the prediction of lead malfunction demonstrated a sensitivity of 91% and a specificity of 92%.

**Figure 3.**
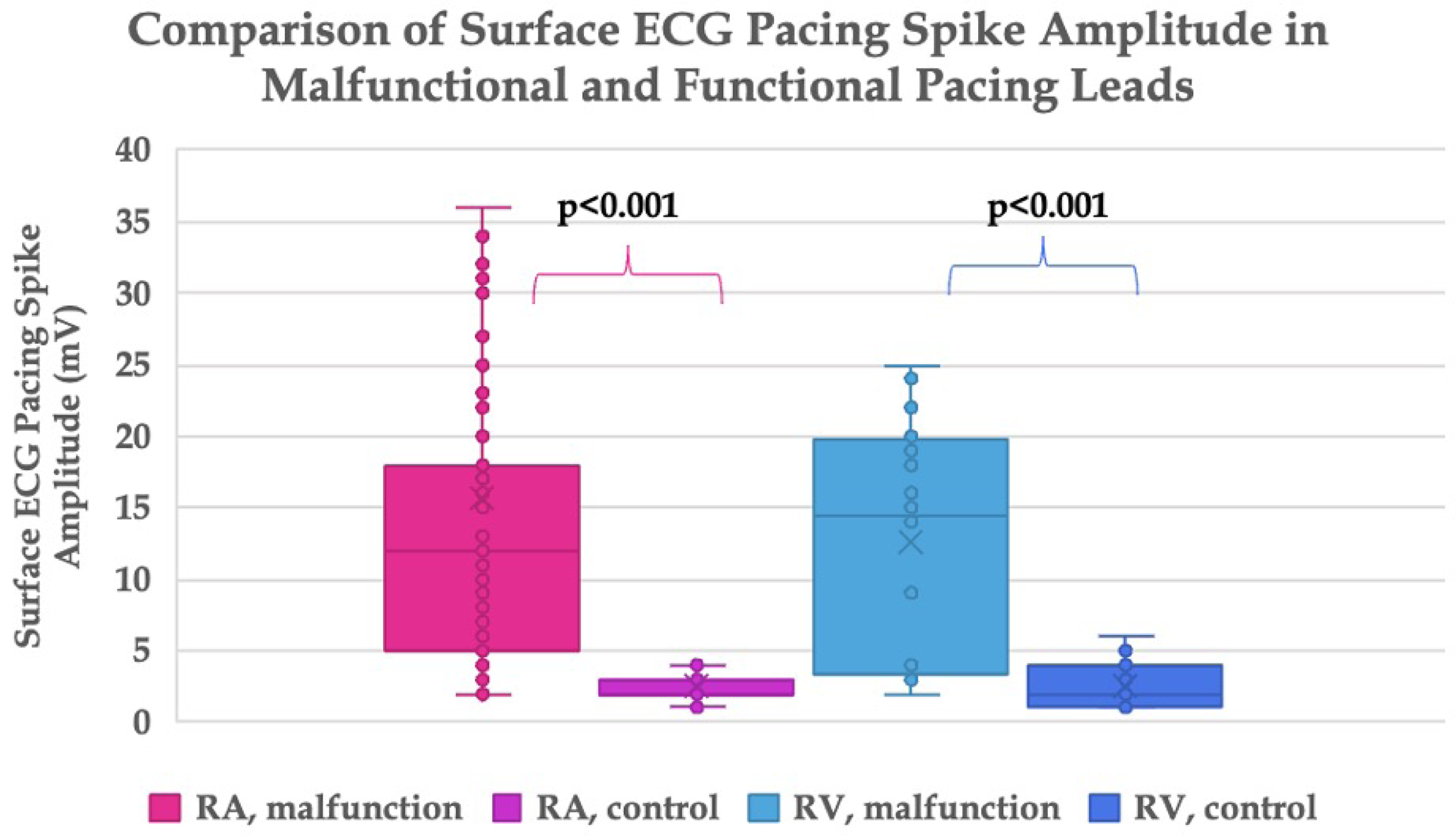
ROC curve for EKG pacing spike absolute amplitude generated for prediction of lead functional status. Accuracy improved when normalized for pacing output, but absolute value curves are shown due to better clinical applicability.

## Discussion

We have demonstrated that a 12 lead ECG measurement of bipolar pacing stimulus amplitude is an accurate means of detecting insulation breach in transvenous CIED leads. To our knowledge, this is the first study to quantify and characterize this finding as well as its clinical utility.

These data are important for several reasons. First, there is an important need for better and adjunctive ways of identifying this type of lead failure. Serious clinical adverse events have been demonstrated by lead malfunction as a result of insulation breach due to pacing inhibition, inappropriate ICD therapy and ineffective delivery of appropriate defibrillator shocks [7] [8]. While this type of lead malfunction commonly manifests as low impedance trends and lead noise detection, impedance is not sensitive to insulation breakdown [3, 6]. In fact, >70% of instances show that lead insulation breakdown is associated with normal impedance measurements [6]. Bipolar stimulus amplitude on ECG would be an adjunct and an easily obtainable variable for this purpose.

Our analysis also demonstrated a large magnitude of effect using this variable when comparing abnormal to normal transvenous leads. The absolute difference between the two groups for both atrial and ventricular leads was approximately 12mV, which in a normally standardized ECG is over 2 large boxes – enabling readily visible differences without the need of digital calipers or other measuring equipment. Using a cutoff amplitude of 5 mm per V allowed for both surprisingly high sensitivity and specificity for detection of this type of lead malfunction. Although a more practical value may be an absolute ECG stimulus amplitude cutoff of 3.5mm, since this does not require knowledge of programmed CIED output. Finally, this parameter is clinically easy to obtain as it does not require device interrogation equipment and is able to be acquired in most clinical settings.

Our data agreed with prior published literature demonstrating pacing impedance as an imperfect variable for identification of this type of lead malfunction. In our dataset we observed no significant differences in lead impedance among normal and malfunctioning RV leads. Additionally, the modestly lower impedance observed in abnormal atrial leads were still within normal range.

Despite our observations, we acknowledge the resolution of a basic surface 12 lead to detect pacemaker stimuli is not high. At a minimum, the sampling frequency of a recording system should be at least twice as fast as the signal being sampled. In this case, a 4000hz sampling rate is 4 times that of a 1ms pacing stimulus, but only 1.6 times faster than that of a stimulus that lasts 0.4ms. The shape and frequency content of the stimulus may vary so that the peak voltage may not be wholly captured on the ECG tracing. Very high stimulus amplitudes are clipped or overlap adjacent lead recordings in standard ECGs so peak amplitudes are likely to have an upper limit for recording.

We termed the proposed mechanism of our findings “pseudo-unipolarization”. It is a known phenomenon that unipolar pacing generates larger amplitude pacing spikes on every ECG lead, while bipolar pacing spikes generate much smaller amplitudes that may not be visible on every channel of the 12-lead ECG [9]. When lead insulation breaks down, conductors come into low resistance contact with the surrounding tissue and offer a parallel current path for a given pacing stimulus. The relative difference in impedances and increased distance between the electrodes between the parallel circuits (the normal ring-tip bipolar and abnormal path created from breakdown) is what accounts for ECG stimulus height recorded on ECG. While only a small portion of this current is diverted through the body’s tissues, the original amplitude of the stimulus is orders of magnitude higher than intrinsic cardiac signal and would thus be able to be detected on 12 lead ECG amplifiers.

## Limitations

Limitations of this study include the inherent shortcomings of its retrospective nature and prospective validation is thus required. Our analysis group included a cohort of patients with known lead malfunction which may have included other abnormalities beyond insulation breach and confounded our results. The gold standard for our experimental group were identified by evidence of exposed conductors to tissue and thus may only represent an advanced subset of insulation breaches. How our parameter behaves in less manifest insulation breaches is not known. Finally, our control group could not be definitively proven to have the complete absence of lead malfunction but was inferred since they were newly implanted.

## Conclusion

Surface ECG bipolar pacing stimulus amplitude holds promise as a means of detecting lead insulation breakdown. Due to the concept of pseudo-unipolarization, the amplitude of a bipolar pacing stimulus on ECG is significantly larger in many malfunctioning leads compared to normal functioning leads in implantable cardiac devices.

## Data Availability

All data can be made available upon request

## Acknowledgements

We would like to acknowledge Cyrus Lloyd for advice on electrical parameters.

